# Distinct patterns of SARS-CoV-2 transmission in two nearby communities in Wisconsin, USA

**DOI:** 10.1101/2020.07.09.20149104

**Authors:** Gage K. Moreno, Katarina M. Braun, Kasen K. Riemersma, Michael A. Martin, Peter J. Halfmann, Chelsea M Crooks, Trent Prall, David Baker, John J. Baczenas, Anna S. Heffron, Mitchell Ramuta, Manjeet Khubbar, Andrea M. Weiler, Molly A. Accola, William M Rehrauer, Shelby L. O’Connor, Nasia Safdar, Caitlin S. Pepperell, Trivikram Dasu, Sanjib Bhattacharyya, Yoshihiro Kawaoka, Katia Koelle, David H. O’Connor, Thomas C. Friedrich

## Abstract

Evidence-based public health approaches that minimize the introduction and spread of new SARS-CoV-2 transmission clusters are urgently needed in the United States and other countries struggling with expanding epidemics. Here we analyze 247 full-genome SARS-CoV-2 sequences from two nearby communities in Wisconsin, USA, and find surprisingly distinct patterns of viral spread. Dane County had the 12th known introduction of SARS-CoV-2 in the United States, but this did not lead to descendant community spread. Instead, the Dane County outbreak was seeded by multiple later introductions, followed by limited community spread. In contrast, relatively few introductions in Milwaukee County led to extensive community spread. We present evidence for reduced viral spread in both counties, and limited viral transmission between counties, following the statewide “Safer at Home” public health order, which went into effect 25 March 2020. Our results suggest that early containment efforts suppressed the spread of SARS-CoV-2 within Wisconsin.

## Introduction

The earliest outbreaks of severe acute respiratory syndrome coronavirus 2 (SARS-CoV-2) in the United States were seeded by travelers who became infected abroad and initiated chains of community transmission. Several months later, SARS-CoV-2 is now ubiquitous. More than 96% of the 3,144 United States administrative subdivisions (i.e., counties, boroughs, and parishes) have reported at least one SARS-CoV-2 case by June 23, 2020 ^1^. Movement between administrative subdivisions and states, rather than introduction from abroad, now poses the greatest risk for seeding new clusters of community transmission. Is it still possible to interrupt the spread of SARS-CoV-2 between nearby counties once community transmission is established?

Case counts from diagnostic SARS-CoV-2 testing are used to understand community transmission, but community-level testing may not be widely available and passive surveillance is unlikely to detect asymptomatic or presymptomatic infections. Viral genome sequencing has emerged as a critical tool to overcome these limitations and provides a complementary means of understanding viral transmission dynamics. The value of this approach was demonstrated during the West African Ebolavirus outbreak in 2014-2016 and again during the emergence of Zika virus in the Americas in 2015-2016 ^2,3^.

The collective global effort to sequence SARS-CoV-2 dwarfs these earlier efforts. As of 28 June 2020, more than 55,000 SARS-CoV-2 sequences collected from over 82 countries have been sequenced and shared publicly on repositories like the Global Initiative on Sharing All Influenza Data (GISAID), enabling real-time phylogenetic analyses encompassing global SARS-CoV-2 diversity ^4^. Patterns of viral sequence variation can also be used to estimate epidemiological parameters, including the total number of infections in a given population and epidemic doubling time, independent of case counts ^4–14^. Here we apply these methods to gain a nuanced view of SARS-CoV-2 transmission within and between regions of the American Upper Midwest.

Dane and Milwaukee Counties are the two most populous counties in the US state of Wisconsin. They are separated by approximately 100 kilometers of rural and suburban communities in Jefferson and Waukesha Counties. An interstate highway that typically carries ~40,000 vehicles a day connects all four of these counties ^15^. Madison and Milwaukee are the largest cities in Wisconsin as well as in Dane and Milwaukee Counties, respectively, and are demographically dissimilar ^16,17^. On 25 March 2020, the Wisconsin Department of Health Services ordered most individuals to stay at home, closed non-essential businesses, and prohibited most gatherings, an order termed “Safer at Home” ^18–20^. While there were some policies enacted to reduce the viral spread prior to this order ^21^, the “Safer at Home” order represented the most significant restriction on individuals and businesses. This Executive Order remained in effect until 13 May 2020, when it was struck down by the Wisconsin Supreme Court. From the start of the Executive Order through 21 April 2020, Dane and Milwaukee Counties had the highest documented number of SARS-CoV-2 cases in Wisconsin. Therefore, these two counties provide a “natural experiment” to understand the impact of the “Safer at Home” Executive Order on within- and between-county SARS-CoV-2 transmission in two nearby US counties with distinguishing demographic features.

Our analyses indicate that the Dane and Milwaukee County SARS-CoV-2 outbreaks were seeded by a different number of introductions and subsequently defined by distinct patterns of viral spread. Despite growing cumulative case counts in both counties, virus transmission clusters remained largely localized within individual counties with evidence of little mixing across counties. Moreover, we find that the virus’s basic reproductive number decreased in both counties evaluated during the time in which the “Safer at Home” order was in place, consistent with adoption of physical distancing, use of face coverings, and other related practices ^22^.

## Results

### SARS-CoV-2 epidemics and community demographics in Dane and Milwaukee Counties

Dane County and Milwaukee County are both located in Southern Wisconsin. Milwaukee County is 127 km east of Dane County, measured from center to center. As of 2015, Dane County had a population of 516,850 at a density of 166 people per km^2^ compared to 952,150 at 1,522 per km^2^ for Milwaukee County (**Fig 1A**) ^16,17^.

**Figure 1.**
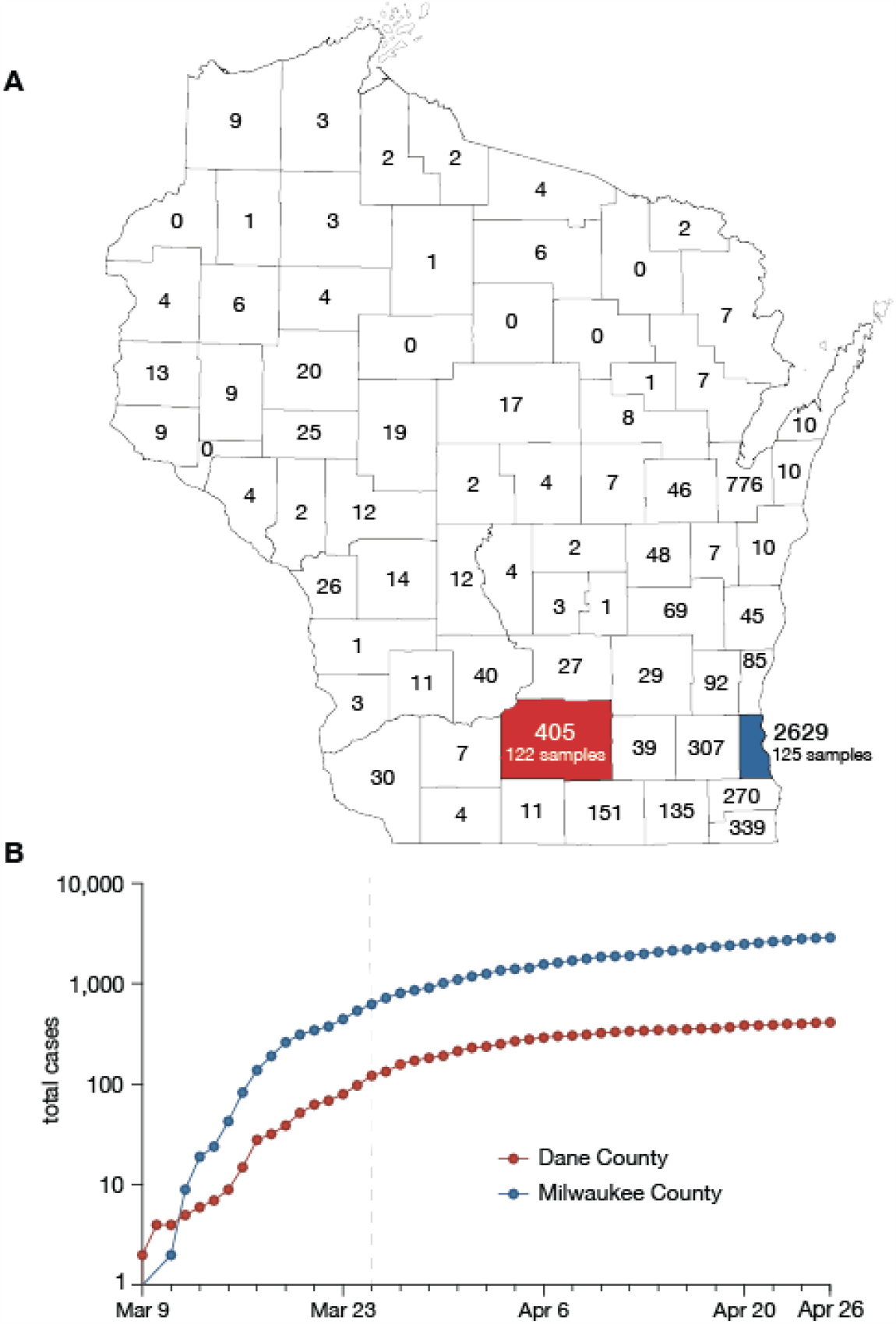
Demography and epidemiology of SARS-CoV-2 in southern Wisconsin. A) A map of Wisconsin highlighting Dane County (red) and Milwaukee County (blue). Cumulative case counts through 26 April 2020 are reported within each county border. B) Cumulative SARS-CoV-2 cases in Dane County (red) and Milwaukee County (blue) from 9 March through 26 April. The vertical dashed line indicates the start date of Wisconsin’s “Safer at Home” order, which went into effect 25 March 2020 ^22^.

The majority of individuals living in Dane County are White (81.5%). The next largest group identifies as Hispanic or Latinx (6.3%), followed by Asian (6.0%), Black (5.9%), and American Indian (0.3%) ^17^. Milwaukee County is less predominantly White (53.3%) with much larger Black (27.2%) and Hispanic or Latinx (14.5%) populations, followed by Asian (4.3%) and American Indian (0.7%) ^16^. The percent of individuals ≥65 years old is similar in Dane County (13.7%) and Milwaukee County (13.6%), while the percent of individuals under 18 years is slightly lower in Dane County (20.4%) than Milwaukee County (24%). In addition, median income and access to healthcare resources is lower in Milwaukee County than in Dane County 23. The median individual in Milwaukee County is also more likely to experience poverty and to live with comorbidities such as type II diabetes, hypertension, and obesity (**Table 1**) ^23^.

**Table 1.** County level demographics for Dane and Milwaukee County.

| County-level demographic data | Dane | Milwaukee |
| --- | --- | --- |
| Population size (2015) | 516,850 | 952,150 |
| Population per square mile (2015) | 430 | 3942 |
| Average number of persons per dwelling (2014-2018) | 2.35 | 2.44 |
| Age (2014-2018): |  |  |
| % of population under 5 | 5.6 | 6.9 |
| % of population under 18 | 20.4 | 24 |
| % of population over 65 | 13.7 | 13.6 |
| Race/ethnicity (2015): |  |  |
| White | 81.5% | 53.3% |
| African American | 5.9% | 27.2% |
| American Indian | 0.3% | 0.7% |
| Hispanic | 6.3% | 14.5% |
| Asian | 6.0% | 4.3% |
| Median income (2015) | \$65,416 | \$45,905 |
| % of population that is uninsured, under 65 (2014-2018) | 4.9% | 8.2% |
| Poverty estimate, all ages (2015) | 11.2% | 20.3% |
| % of population reported overweight or obese (2012-2016) | 54.3% - 58.5% | 64.7% - 69% |
| % of adults reporting diagnosed diabetes (2012-2016) | 4.2% - 6.8% | 8.6% - 9.8% |

Dane County is home to the 12th reported SARS-CoV-2 case in the United States, detected on 30 January 2020. Subsequent cases were not reported until 9 March 2020. By 26 April, Dane County had 405 confirmed SARS-CoV-2 cases and 19 deaths ^24^. Milwaukee County reported its first case on 11 March 2020. By 26 April, Milwaukee County had reported 2,629 confirmed SARS-CoV-2 infections and 126 deaths ^25^ (**Fig 1B**).

Sequences for this study were derived from 247 nasopharyngeal (NP) swab samples collected from Dane County between 14 March 2020 through 18 April 2020, and Milwaukee County from 12 March 2020 though 26 April 2020, Wisconsin. Additional sample metadata are available in **Supplemental Information 1**.

### Dane and Milwaukee County viruses are genetically distinct

If an outbreak is fueled by community spread following a single introduction, one would expect viral genomes to be close phylogenetic relatives, in which case genetic distances measured in any pairwise comparisons of sequences would be low. To examine this, we generated SARS-CoV-2 consensus sequences using the ARTIC Network protocol ^26,27^ and defined the population of consensus single-nucleotide variants (SNVs) relative to the initial SARS-CoV-2 Wuhan reference (Genbank: MN908947.3).

In Dane County, we identified 155 distinct SNVs across 122 samples evaluated. These SNVs are evenly distributed throughout the genome, and 92.9% (144/155) are located in open reading frames (ORFs). In Dane County, 52.9% (82/155) of consensus SNVs result in an amino acid change (nonsynonymous) and 40% (62/155) do not (synonymous) (**Fig 2A**).

**Figure 2.**
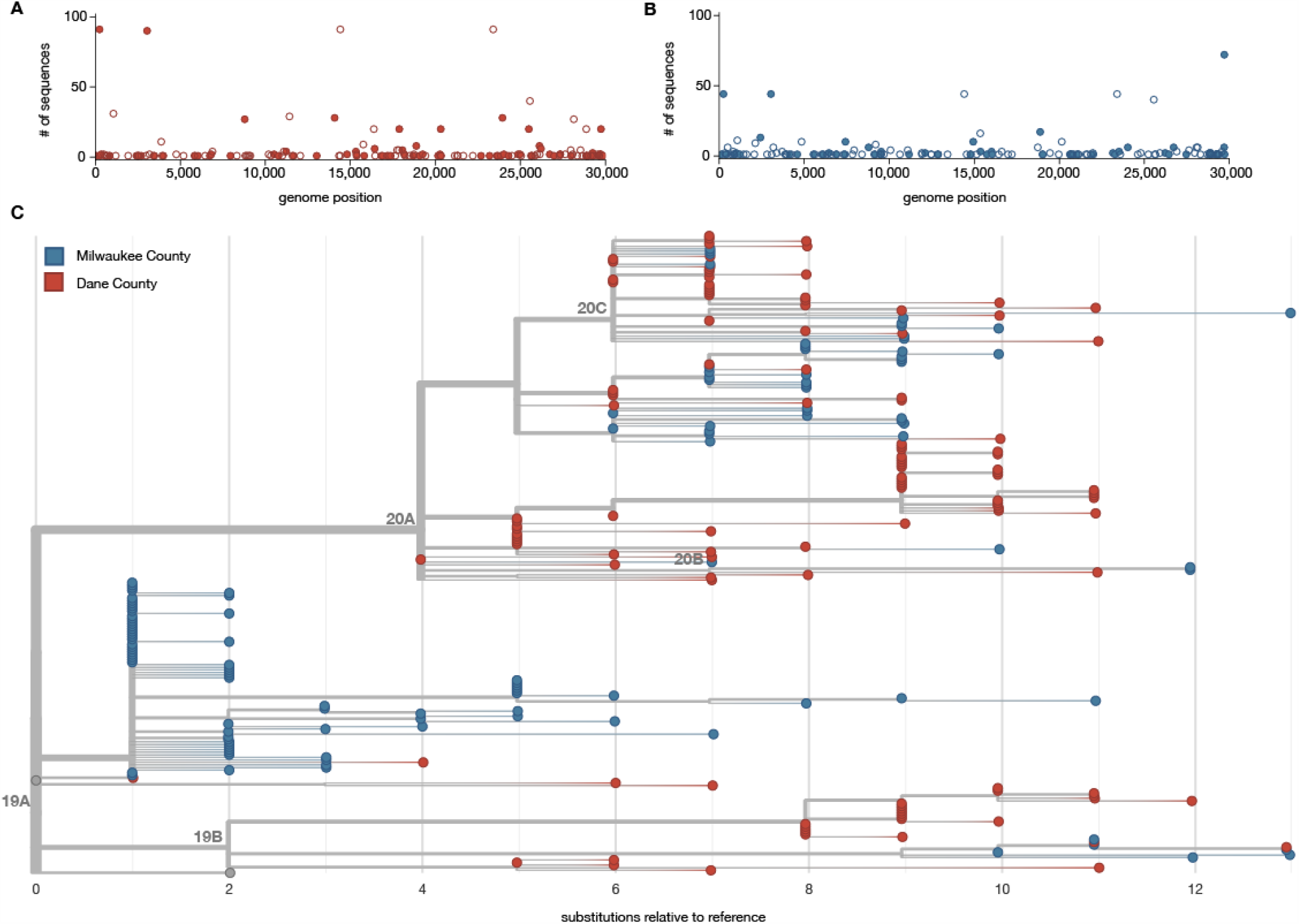
Characterizing consensus-level variants and sequence divergence among Dane and Milwaukee County sequences. SNVs are annotated relative to the initial Wuhan SARS-CoV-2 reference (Genbank: MN908947.3). A) Frequency of consensus SNVs among the Dane County sequences. B) Frequency of consensus SNVs among the Milwaukee County sequences. Open symbols denote synonymous or intergenic SNVs and closed symbols denote nonsynonymous SNVs. C) A divergence-based phylogenetic tree built using Nextstrain tools for the 122 Dane County (red) and 125 Milwaukee County (blue) sequences. Wisconsin samples are rooted against Wuhan-Hu-1/2019 and Wuhan/WH01/2019.

In Milwaukee County, we identified 148 distinct SNVs across 125 samples evaluated. Among the observed consensus SNVs in Milwaukee County, 63.5% (94/148) are nonsynonymous and 31.8% (47/148) are synonymous (**Fig 2B**).

Mean inter-sequence pairwise SNV distance was 7.65 (std 1.83) and 5.02 (std 3.63) among Dane County and Milwaukee County sequences, respectively (**Fig 2C**). Likewise, we detected an average of 4.4 new SNVs per day (sampling period of 35 days) in Dane County and 3.6 new SNVs per day (sampling period of 41 days) in Milwaukee County. Previous reports suggested SARS-CoV-2 is expected to acquire approximately one fixed SNV every fifteen days following a single introduction ^28^. Compared to this benchmark, both Dane County and Milwaukee County have “excess” diversity which can be most parsimoniously explained by multiple introductions of divergent viruses. These patterns are consistent with a greater number of introductions of distinct viruses into Dane County compared to Milwaukee County.

To further analyze genetic differences among viruses in the two locations, we assigned clades using the Nextstrain nomenclature. For example, clade 19B is defined by two mutations at nucleotides 8,782 (ORF1ab S2839S) and 28,144 (Spike L84S) relative to a reference SARS-CoV-2 isolate from Wuhan, China (Genbank: MN908947.3). The majority of Dane County sequences, 51.6% (n = 63 sequences), cluster in the 20A clade (**Fig 3A**). This clade is defined by four variants, at nucleotide positions 241 (upstream of the first open reading frame), 3,037 (ORF1a F924F), 14,408 (ORF1b P314L), and 23,403 (S D614G). A minority (n = 31 sequences; 24.8%) of Milwaukee County sequences also cluster in this clade. In contrast, the 19A clade is most common (n = 75 sequences; 60.0%) in sequences from Milwaukee County. This clade is distinguished by a U-to-C variant at nucleotide position 29,711 (downstream of ORF10) (**Fig 3B**).

**Figure 3.**
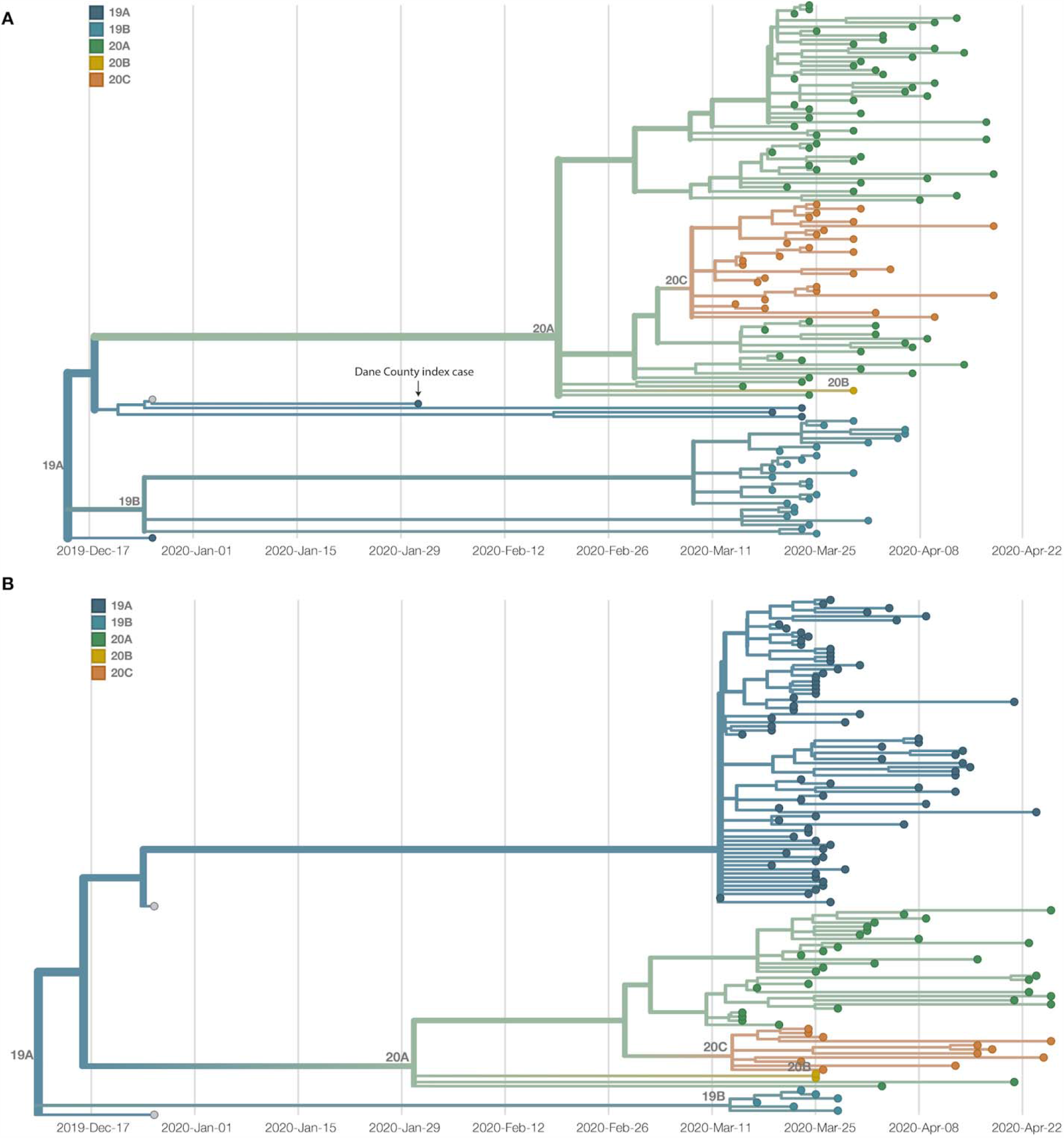
Dane and Milwaukee County outbreaks are defined by genetically distinct viruses. A) A time-resolved phylogenetic tree built using Nextstrain tools for 122 samples collected in Dane County. B) A time-resolved phylogenetic tree for 125 samples collected in Milwaukee County. Clade is denoted by color. Both phylogenies include Wuhan sequences (Wuhan-Hu-1/2019 and Wuhan/WH01/2019, denoted in grey) to more effectively time-align each tree.

### No onward spread from Dane County index case

The first known SARS-CoV-2 case in Wisconsin was a person who was likely infected during travel to Wuhan, Hubei province, China, where they were exposed to family members with confirmed SARS-CoV-2 infections. The patient reported a sore throat shortly before departing China and returning to the US on 30 January 2020. The person wore a mask during the return flight. Upon arrival in the US, the person immediately presented to an emergency department while still wearing a mask. The person was afebrile and had no respiratory or gastrointestinal signs or symptoms, but began to develop mild respiratory symptoms shortly thereafter. The person’s condition remained stable and never required hospitalization or advanced care, with symptoms resolving five days later. The person self-quarantined in an isolated room in a home with a dedicated bathroom for 30 days following symptom onset. During this time, nasopharynx samples repeatedly tested positive for SARS-CoV-2 viral RNA.

Documentation of asymptomatic infections of SARS-CoV-2 has led to concerns about the role of cryptic community transmission in the United States ^7,29,30^. However, sequencing in other locations in the United States has revealed early introduction events did not always go on to seed downstream community spread ^31^. To determine whether SARS-CoV-2 cases detected in Dane County in March might have been due to undetected spread from the first Wisconsin introduction, we compared the sequence of this early case with local and global SARS-CoV-2 sequence diversity. The first Dane County patient’s virus contains an in-frame deletion at nucleotide positions 20,298 - 20,300, in a region that codes for the poly(U)-specific endoribonuclease; the impact of this mutation on viral fitness is unknown ^32^ (**Supplemental Fig 1**). Notably, this deletion was not detected in any other Dane County sequence, nor in any other sample(s) submitted to GISAID as of 18 April 2020. Moreover, there are no branches originating from the index Dane County case on either the global (Wisconsin sequences plus a subsampled set of global sequences) or local phylogenies (Wisconsin sequences only, maximum likelihood) (**Fig 2C, Fig 3A**). Thus, this early case appears to be an example of successful infection control practices.

### SARS-CoV-2 outbreak dynamics differ between Milwaukee and Dane Counties

The independent local phylogenies in Dane and Milwaukee County suggested that these two nearby locations had largely distinct SARS-CoV-2 epidemics through April 2020. To better understand the number of introductions and continued transmission dynamics, we generated a time-resolved sub-sampled global phylogeny incorporating Dane County (red tips) and Milwaukee County (blue tips) sequences alongside representative global SARS-CoV-2 sequences, including all other available Wisconsin sequences (purple tips) (**Fig 4A**). Dane County viruses are distributed throughout the tree, consistent with multiple unique introductions. In contrast, Milwaukee County viruses cluster more closely together, consistent with fewer introductions and subsequent community transmission.

**Figure 4.**
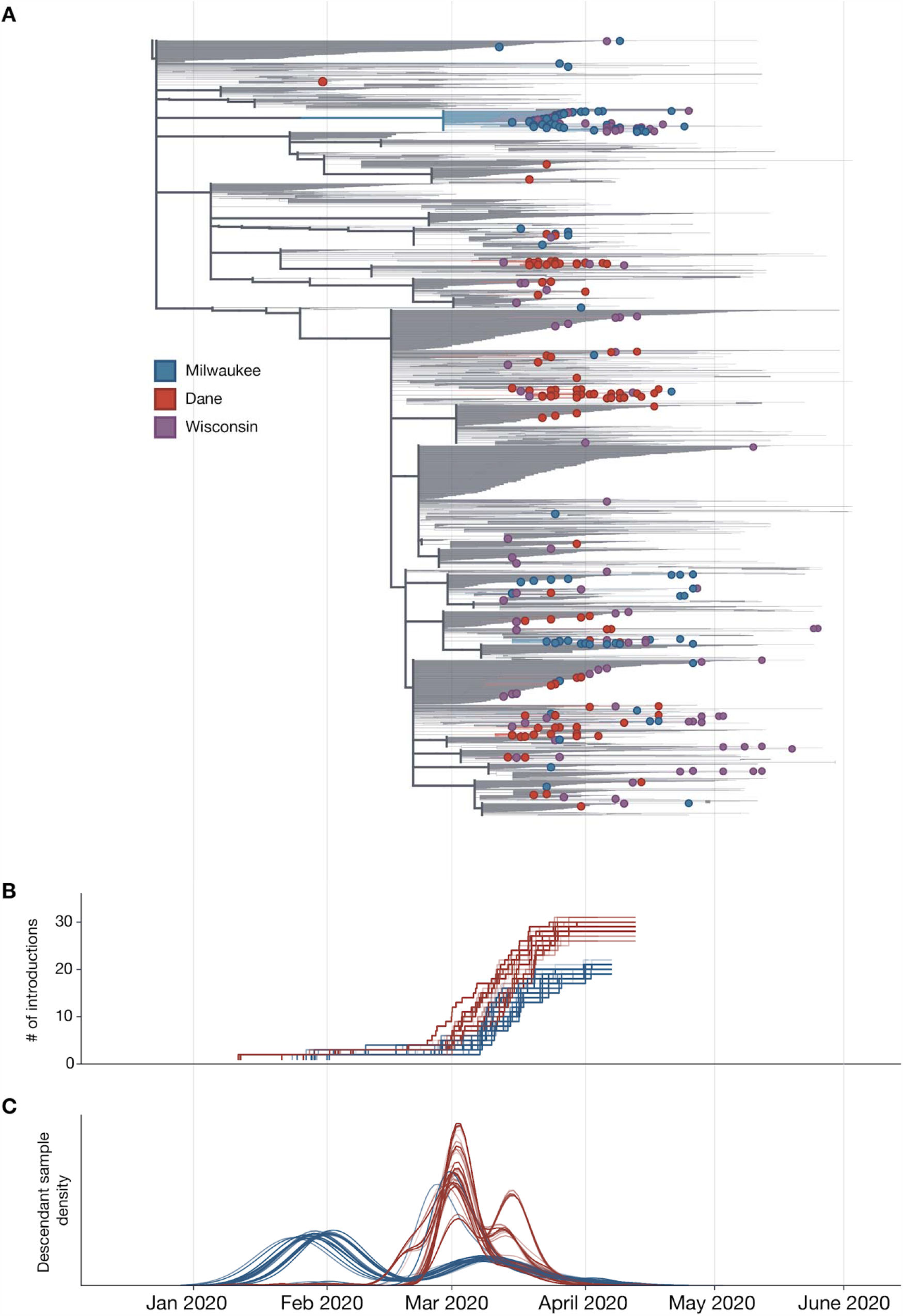
Estimate of the number of introduction events into Milwaukee and Dane County and their relative contribution to downstream epidemic dynamics. A) Maximum likelihood (ML) time-resolved tree with subsampled global sequences and closest phylogenetic neighbors relatives included (grey branches). Sequences from Dane and Milwaukee Counties are highlighted in red and blue, respectively. Sequences with geolocation information available to the state level, or that are located outside of Dane and Milwaukee Counties (i.e. La Crosse) are shown in purple. B) Estimated cumulative number of introduction events into each county. C) Gaussian Kernel Density Estimate plots showing the estimated timing of each introduction event (3 curves per replicate: mean and 90% confidence intervals) into Dane County (red) or Milwaukee County (blue). The relative number of samples from each region attributable to an introduction event is represented on the y-axis. Curves are normalized to a cumulative density of one; therefore, y-axis scale is not shown.

To estimate the number of introductions into the state and subsequently each county, we used an ancestral state reconstruction of internal nodes. We performed 100 bootstrap replicates to account for uncertainty in the phylogenetic inference. This yielded an estimate of 59 [59, 63] (median [95% highest density interval (HDI)]) independent introductions of SARS-CoV-2 into the state of Wisconsin. Of these, 29 [28, 31] led to introductions into Dane county whereas only 21 [19, 21] led to introductions into Milwaukee county (**Fig 4B)**. Surprisingly, only 9 [6, 10] of the introductions into Wisconsin were associated with sequences from both counties. Furthermore, these shared introductions accounted for only 20-30% of the samples from Dane and Milwaukee County present in our dataset. Together, our analyses suggest that transmission between Dane and Milwaukee counties has not been a principal component of viral spread within either region. We find that local transmission in Milwaukee County began earlier, with an introduction event in late January/early February leading to a large number of the Milwaukee County sequences (**Fig 4C**). In comparison, most samples collected from Dane County are associated with multiple introductions in late February/early March (**Fig 4C**). Despite the fact that there were more introductions into Dane County, the reported number of cases was considerably less than in Milwaukee County. This indicates that each introduction into Dane County contributed less to onward viral transmission than in Milwaukee County.

To account for sampling bias on our estimates, we randomly sampled sequences from our set of Dane and Milwaukee County samples (N = 20-240, increments of 20) and pruned all other Dane and Milwaukee samples from the maximum likelihood tree. This was repeated 10 times for each N, creating a set of 120 trees. We repeated the ancestral state reconstruction on each of these trees and re-estimated the number of introductions (**Supplemental Fig 2**). The number of estimated introductions into Dane County continued to increase with the number of sampled sequences, indicating that these data may be undersampling the true circulating viral lineages. In contrast, the number of estimated introductions into Milwaukee County decreases more slowly than Dane County, consistent with a small number of introductions. Although, we cannot rule out that the small number of introductions in Milwaukee County is an artifact of biased sampling, where the available sequences may only represent a portion of the transmission chains and not a true estimation of the total circulating viral population. Because of this, the true number of introductions is likely to change as more sequences become available in each county.

### Spread of SARS-CoV-2 was reduced following Wisconsin’s “Safer at Home” Order

We next used viral sequence data to assess the impact of Wisconsin’s “Safer at Home” order on the basic reproduction number (R_0_). Given the role of superspreading dynamics in SARS-CoV-2 epidemics ^9,33,34^, we evaluated the impact on R_0_ for the Dane County and Milwaukee County epidemics in low, medium, and high transmission heterogeneity scenarios, where the level of transmission heterogeneity reflects the role for superspreading events, i.e high transmission heterogeneity reflects many supersupreading events. In both counties, under all three scenarios, R_0_ fell by at least 40% after 25 March, indicating that the sequencing data support the observed decline in reported cases. In Dane County, estimated median R_0_ was reduced by 40% [4, 74], 49% [13, 79], and 60% [30, 83] under low, medium, and high transmission heterogeneity, respectively. Similarly, in Milwaukee County, estimated median R_0_ was reduced by 68% [50, 83], 71% [56, 86], and 72% [60, 84] under low, medium, and high transmission heterogeneity, respectively.

In Dane County, estimated cumulative incidence was best predicted with the medium transmission heterogeneity model based on alignment with reported incidence (**Fig 5A**). Whereas Milwaukee County’s cumulative incidence was best predicted with the model using high transmission heterogeneity (**Fig 5B**). A greater role for superspreading events in Milwaukee versus Dane County could be explained by higher population density, higher poverty rates, and worse healthcare access (**Table 1**), all of which may increase contact rates and impede physical distancing efforts ^34–38^. Assuming moderate transmission heterogeneity in Dane County, estimated R_0_ prior to 25 March was 2.24 [1.86, 2.65] and the median estimated cumulative incidence at the end of the study period (26 April) was 4,546 infections [1,187, 23,709] compared to 405 positive tests. In contrast, assuming high transmission heterogeneity in Milwaukee County, estimated R_0_ prior to 25 March was 2.82 [2.48, 3.20] and the median cumulative incidence on 26 April was only 3,008 infections [1,483, 7,508] compared to 2,629 positive tests.

**Figure 5.**
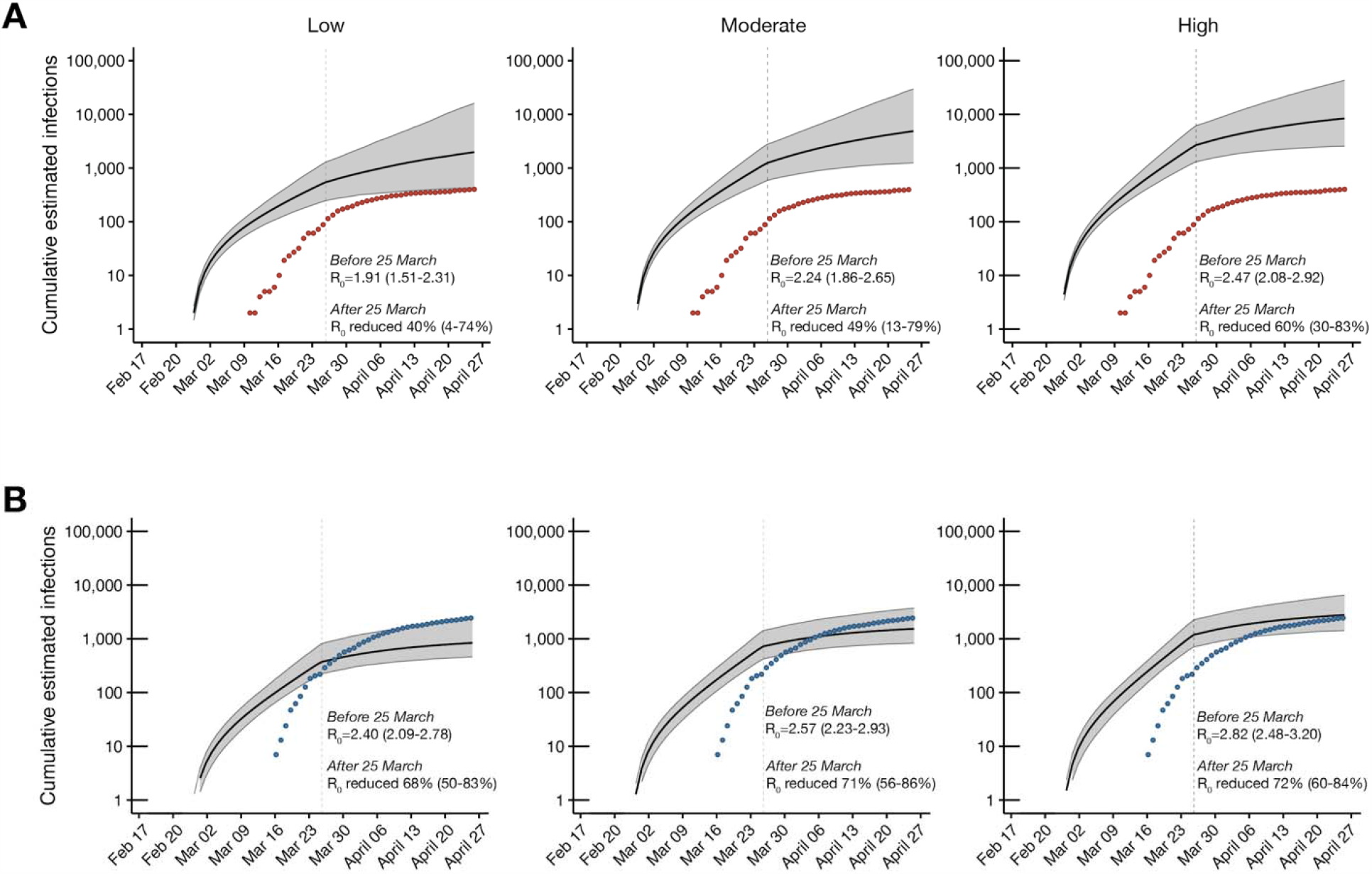
Phylodynamic modelling of regional outbreaks informs regional outbreak dynamics before and after government interventions. Bayesian phylodynamic modelling of cumulative incidence up to 26 April for outbreaks in A) Dane County and B) Milwaukee County under low (left), medium (center), and high (right) transmission heterogeneity conditions. Model parameters for low, medium, and high transmission heterogeneity were fixed such that 20, 10, and 5% of superspreading events contribute 80% of cumulative infections, respectively. Median cumulative incidence (solid black line) is bound by the 95% confidence intervals (CI; gray ribbon). Dots represent reported cumulative positive tests in Dane County (red) and Milwaukee County (blue). Estimated median reproductive numbers (R_0_) with 95% HDI are listed for the period before the Wisconsin “Safer at Home” order was issued on 25 March 2020. Percent reduction in R_0_ with 95% HDI is provided for the period after 25 March 2020.

With passive SARS-CoV-2 surveillance efforts in both counties likely missing subclinical and asymptomatic SARS-CoV-2 infections, we expect the true cumulative incidence to be considerably greater than the reported incidence, as has been suggested by others ^39^. Indeed, estimated cases were ~10x higher than reported cases in Dane County. Given that there were no substantial differences in the surveillance efforts between counties, we expected more than the 1.1-fold difference in estimated and reported cases in Milwaukee County. Nearly equivalent estimated and reported cumulative incidence in Milwaukee County could be explained by better detection rates, inaccurate model parameters, and/or biased sampling. With better detection rates, a greater proportion of actual infections would be reported, but given the similar surveillance efforts between counties we expect detection rates to be comparable. Another possible explanation we cannot rule out is that different model parameters are required to more accurately model Milwaukee County’s epidemic. Our testing of three superspreading scenarios demonstrated that the superspreading parameters, at least, may be county-specific. In the case of biased sampling, where the available sequences only represent a portion of transmission chains in the county, our model would only estimate the caseload resulting from a subset of transmission chains in Milwaukee County and would underestimate the county-wide caseload. In support of representative county-wide sampling in Dane, but not Milwaukee County, sequences from 26.4% (107/405) of test-positive cases in Dane County, but only 3.9% (117/3008) of test-positive cases in Milwaukee County were available for phylodynamic modelling ^24,25^.

## Discussion

Dane County, Wisconsin had one of the earliest detected cases of SARS-CoV-2 infection in the United States, but this infection did not spark community spread. This is probably due to a combination of good infection control practices by healthcare providers, the patient, and sheer luck. Since March 2020 we find evidence for extensive introductions of SARS-CoV-2 into Dane County, none of which led to large-scale transmission clusters by the end of April 2020. As of 24 June 2020, Dane County has had a cumulative prevalence of 233 cases per 100,000 residents. In contrast, Milwaukee County, a larger and more densely populated region ~100km away, has had 1,105 cases per 100,000 residents as of 24 June 2020 ^40^. Our findings suggest that Milwaukee County’s higher caseload stems from greater levels of community spread descendant from fewer introduction points than in Dane County. Strikingly, we see little evidence for mixing of virus populations between these two closely-linked communities in the same US state.

We used patterns of SARS-CoV-2 diversification in a phylodynamic model to estimate the initial reproductive rate of infections in each county before official social distancing policies were enacted. In this initial phase of the outbreak, the median estimated R_0_ trended lower in Dane County than in Milwaukee County (2.24 vs 2.82). Higher population density in Milwaukee County could have contributed to a higher reproductive rate. A potential additional explanation for greater community spread in Milwaukee County is that the average individual in Milwaukee County, compared to Dane County, has access to fewer financial and healthcare resources and is more likely to experience poverty and to live with comorbid conditions, many of which are also risk factors for testing positive for SARS-CoV-2, the latter of which are also risk factors for severe Coronavirus Disease (COVID-19) ^16,17,41,42^. Additionally, Milwaukee County is home to a higher proportion of Black and Hispanic or Latinx individuals compared to Dane County.

Because of race-based discrimination, people belonging to these groups experience worse health outcomes than White individuals, despite being treated in the same healthcare systems ^16,17,43,44^. The social vulnerability index (SVI) is a metric ranging designed to determine how resilient a community is when confronted with external stressors like natural disasters or a pandemic ^45^. A higher SVI indicates a community is vulnerable to experiencing worsened outcomes secondary to an external stressor (range of zero to one). All of the factors mentioned above contribute to a higher SVI in Milwaukee County (0.8268) compared to Dane County (0.1974) ^45^. While the association between SIV and SARS-CoV-2 indicidence is not significant, according to a recent study, the SVI sub-components of socioeconomic and minority status are both predictors of higher SARS-CoV-2 incidence and case fatality rates ^46^. These sub-components are likely to be among the main drivers in the outbreak dynamics between Dane and Milwaukee County.

Like most US states, in late March 2020 Wisconsin enacted a set of social distancing policies aimed at reducing the spread of SARS-CoV-2. Wisconsin’s order, termed “Safer at Home,” was enacted 25 March 2020. After this time point, the estimated R_0_ was reduced by 40% or more in both counties. The sequencing data is consistent with the observed reduction in positive tests, as clusters expanded more slowly and new clusters arose more slowly. Throughout this time, we find that the Dane County and Milwaukee County outbreaks were largely independent of one another. Our data reveal only limited mixing of SARS-CoV-2 genotypes between these geographically-linked communities, supporting the notion that public health policies emphasizing physical distancing effectively reduce transmission between communities. Notably, “Safer at Home” ended abruptly 13 May 2020, when it was overturned by the Wisconsin Supreme Court. Additional sequencing and epidemiological data will be necessary to understand whether virus intermingling between these counties increased after the cessation of the Executive Order.

Viral determinants could also affect differential transmission patterns within and between Dane and Milwaukee Counties. If variants with greater transmission potential exist, then early introductions of such a variant into a community could contribute to greater spread there. Recent reports have suggested that a point mutation in the SARS-CoV-2 spike protein-encoding an aspartate-to-glycine substitution at amino acid residue 614 (D164G) may enhance transmissibility. This mutation confers increased infectivity of pseudotyped murine retroviruses in ACE2-expressing HEK293T cells ^47^ and has been proposed to be increasing in global prevalence, perhaps under natural selection ^48^. Importantly, however, the rise in D614G frequency could also be due to founder effects, as viruses bearing the glycine allele may have been the first to establish local transmission in Europe. D614G is one of the mutations defining the 20A clade; these viruses remain dominant in Europe ^31^, so introductions from Europe into the United States, including into Dane County, predominantly carry D614G. In comparison, in Milwaukee County, the vast majority of viruses have an aspartic acid residue at this site despite much higher levels of community transmission. This observation does not necessarily indicate that D614G does not impact viral transmissibility; its role may be muted by other determinants of transmission, including demographic and socioeconomic factors.

There are some important caveats to this study. Of the total reported positives in each county during the study period, high-quality sequences were available for 27% of test-positive cases in Dane County, but only 5% of test-positive cases in Milwaukee County ^24,25^. Despite the deep sampling of SARS-CoV-2 sequences in Wisconsin relative to other regions in the US, even greater targeted sequencing efforts may be required to fully capture the sequence heterogeneity conferred by multiple introduction events and variable superspreading dynamics. It is possible additional sequencing in Milwaukee County would uncover additional viral lineages, or that the 5% of cases we sequenced do not fully represent the diversity of viruses found throughout the county, skewing our observations. However, in analyzing sample metadata we find no evidence that particular locations within Milwaukee County were over- or under-sampled relative to their known SARS-CoV-2 prevalence. Another potential explanation is that Milwaukee County was under-testing their epidemic. Throughout the period analyzed here, the percentage of SARS-CoV-2 tests returning positive in Milwaukee County was ~20%, compared to only ~5% in Dane County ^24,25^. As we are only able to sequence test-positive samples, under-testing in Milwaukee County may have limited our ability to capture a complete representation of their epidemic. Through increased testing and continued sequencing efforts, it is likely that we will be able to more fully understand the Milwaukee County outbreak.

It is also possible that other sequences from these counties relevant to our analyses were collected by other groups. As of 21 June 2020, there were 477 Wisconsin sequences available, but only 351 of these had geolocation information resolved to the county level. Some of the remaining 126 sequences likely originated from Dane County or Milwaukee County, but we cannot include these sequences in our analysis given their geolocation data resolved only to the state level. Currently there is no clearly stated national-level guidance for metadata to be associated with pathogen sequences. Dates and geographic locations with greater than state-level resolution are required to track the emergence and spread of novel pathogens like SARS-CoV-2. Explicit regulatory guidance from the United States enabling the disclosure of sequencing data with county-level geolocation data and sampling dates would enable other institutions to harmonize reporting of viral sequences and improve subsequent studies comparing viral sequences from different locations. Such reporting may be especially important for identifying disparities in viral transmission due to socioeconomic vulnerabilities in specific counties that would otherwise be masked using state-level data reporting.

Here we provide the first insights into the emergence and spread of SARS-CoV-2 in Southern Wisconsin. We show an early introduction of SARS-CoV-2 that did not go on to seed downstream community spread. European lineages account for multiple later introductions in Dane County, but we find little evidence for large-scale community spread stemming from any single introduction. Conversely, SARS-CoV-2 lineages from Asia account for relatively fewer unique introductions into Milwaukee County and are followed by increased community spread. We show strong evidence for a reduction in case counts in both Dane and Milwaukee Counties following implementation of Wisconsin’s state-wide “Safer at Home” order, emphasizing the ongoing importance of physical distancing and limiting large gatherings, especially in spaces with limited airflow ^49^. The factors contributing to greater community transmission in Milwaukee County and extinction of infection clusters within Dane County remain unclear, but regional demographics likely play a critical role in these differences. To this end, continued efforts to sequence SARS-CoV-2 viruses across multiple spatio-temporal scales remain critical for tracking viral transmission dynamics within and between communities and for guiding “precision medicine” public health interventions to suppress future SARS-CoV-2 outbreaks.

## Methods

### Sample approvals and sample selection criteria

Work with residual diagnostic specimens was performed at biosafety level-3 containment at the AIDS Vaccine Research Laboratory at the University of Wisconsin – Madison. Waiver of HIPAA Authorization and approval to obtain the clinical samples along with a Limited Data Set was provided by the Western Institutional Review Board (WIRB #1-1290953-1).

### County-level case data and demographics

The county level map of Wisconsin was obtained from the State Cartographer’s Office (https://www.sco.wisc.edu/maps/wisconsin-outline/). Wisconsin county-level COVID-19 cumulative case data was obtained from the Wisconsin Department of Health Services COVID-19 dashboard (https://data.dhsgis.wi.gov/datasets/covid-19-historical-data-table/, https://cityofmadison.maps.arcgis.com/apps/opsdashboard/index.html#/e22f5ba4f1f94e0bb0b9529dc82db6a3, and https://county.milwaukee.gov/EN/COVID-19). All Dane and Milwaukee county demographic data came from the Wisconsin Department of Health Services Data & Statistics (https://www.dhs.wisconsin.gov/stats) or the U.S. Census Bureau QuickFacts table (https://www.census.gov/quickfacts/fact/table/).

### vRNA isolation

Nasopharyngeal swabs received in transport medium (VTM) were briefly centrifuged at 14,000 r.p.m. for 30 seconds at room temperature to ensure all residual sample sediments at the bottom of the tube. Viral RNA (vRNA) was extracted from 100 μl of VTM using the Viral Total Nucleic Acid Purification kit (Promega, Madison, WI, USA) on a Maxwell RSC 48 instrument and was eluted in 50 μL of nuclease free H2O.

### vRNA isolation for index Dane County Sample

Approximately 140 µL of VTM was passed through a 0.22µm filter (Dot Scientific, Burton, MI, USA). Total nucleic acid was extracted using the Qiagen QIAamp Viral RNA Mini Kit (Qiagen, Hilden, Germany), substituting carrier RNA with linear polyacrylamide (Invitrogen, Carlsbad, CA, USA) and eluting in 30 µL of nuclease free H_2_O. The sample was treated with TURBO DNase (Thermo Fisher Scientific, Waltham, MA, USA) at 37°C for 30 min and concentrated to 8µL using the RNA Clean & Concentrator-5 kit (Zymo Research, Irvine, CA, USA). The full protocol for nucleic acid extraction and subsequent cDNA generation is available at https://www.protocols.io/view/sequence-independent-single-primer-amplification-o-bckxiuxn.

### Complementary DNA (cDNA) generation

Complementary DNA (cDNA) was synthesized using a modified ARTIC Network approach ^26,27^. Briefly, vRNA was reverse transcribed with SuperScript IV Reverse Transcriptase (Invitrogen, Carlsbad, CA, USA) using random hexamers and dNTPs. Reaction conditions were as follows: 1μL of random hexamers and 1µL of dNTPs were added to 11 μL of sample RNA, heated to 65°C for 5 minutes, then cooled to 4°C for 1 minute. Then 7 μL of a master mix (4 μL 5x RT buffer, 1 μL 0.1M DTT, 1µL RNaseOUT RNase Inhibitor, and 1 μL SSIV RT) was added and incubated at 42°C for 10 minutes, 70°C for 10 minutes, and then 4°C for 1 minute.

### Complementary DNA (cDNA) generation for index Dane County sample

Complementary DNA (cDNA) was synthesized using a modified Sequence Independent Single Primer Amplification (SISPA) approach described by Kafetzopoulou et al. ^50,51^. RNA was reverse-transcribed with SuperScript IV Reverse Transcriptase (Invitrogen, Carlsbad, CA, USA) using Primer A (5’-GTT TCC CAC TGG AGG ATA-(N_9_)-3’). Reaction conditions were as follows: 1µL of primer A was added to 4 µL of sample RNA, heated to 65°C for 5 minutes, then cooled to 4 °C for 5 minutes. Then 5 µL of a master mix (2 μL 5x RT buffer, 1 μL 10 mM dNTP, 1 μL nuclease free H_2_O, 0.5 μL 0.1M DTT, and 0.5 μL SSIV RT) was added and incubated at 42°C for 10 minutes. For generation of second strand cDNA, 5 µL of Sequenase reaction mix (1 μL 5x Sequenase reaction buffer, 3.85 μL nuclease free H_2_O, 0.15 μL Sequenase enzyme) was added to the reaction mix and incubated at 37°C for 8 minutes. This was followed by the addition of a secondary Sequenase reaction mix (0.45 μl Sequenase Dilution Buffer, 0.15 μl Sequenase Enzyme), and another incubation at 37°C for 8 minutes. Following incubation, 1µL of RNase H (New England BioLabs, Ipswich, MA, USA) was added to the reaction and incubated at 37°C for 20 min. Conditions for amplifying Primer-A labeled cDNA were as follows: 5 µL of primer-A labeled cDNA was added to 45 µL of AccuTaq master mix per sample (5 µL AccuTaq LA 10x Buffer, 2.5 µL dNTP mix, 1µL DMSO, 0.5 µL AccuTaq LA DNA Polymerase, 35 µL nuclease free water, and 1 µL Primer B (5′-GTT TCC CAC TGG AGG ATA-3′). Reaction conditions for the PCR were: 98°C for 30s, 30 cycles of 94°C for 15 s, 50°C for 20 s, and 68°C for 2 min, followed by 68°C for 10 min.

### Multiplex PCR to generate SARS-CoV-2 genomes

A SARS-CoV-2-specific multiplex PCR for Nanopore sequencing was performed, similar to amplicon-based approaches as previously described ^26,27^. In short, primers for 96 overlapping amplicons spanning the entire genome with amplicon lengths of 500bp and overlapping by 75 to 100bp between the different amplicons were used to generate cDNA. cDNA (2.5 μL) was amplified in two multiplexed PCR reactions using Q5 Hot-Start DNA High-fidelity Polymerase (New England Biolabs, Ipswich, MA, USA) using the conditions previously described ^26,27^.

Samples were amplified through 25 cycles of PCR and each resulting multiplex sample was pooled together before ONT library prep.

### Library preparation and sequencing

Amplified PCR product was purified using a 1:1 concentration of AMPure XP beads (Beckman Coulter, Brea, CA, USA) and eluted in 30μL of water. PCR products were quantified using Qubit dsDNA high-sensitivity kit (Invitrogen, USA) and were diluted to a final concentration of 1 ng/μl. A total of 5ng for each sample was then made compatible for deep sequencing using the one-pot native ligation protocol with Oxford Nanopore kit SQK-LSK109 and its Native Barcodes (EXP-NBD104 and EXP-NBD114) ^27^. Specifically, samples were end repaired using the NEBNext Ultra II End Repair/dA-Tailing Module (New England Biolabs, Ipswich, MA, USA).

Samples were then barcoded using 2.5µL of ONT Native Barcodes and the Ultra II End Repair Module. After barcoding, samples were pooled directly into a 1:1 concentration of AMPure XP beads (Beckman Coulter, Brea, CA, USA) and eluted in 30µL of water. Samples were then tagged with ONT sequencing adaptors according to the modified one-pot ligation protocol ^27^. Up to 24 samples were pooled prior to being run on the appropriate flow cell (FLO-MIN106) using the 72hr run script.

### Processing raw ONT data

Sequencing data was processed using the ARTIC bioinformatics pipeline (https://github.com/artic-network/artic-ncov2019), with a few modifications. Briefly, we have modified the ARTIC pipeline so that it demultiplexes raw fastq files using qcat as each fastq file is generated by the GridION (https://github.com/nanoporetech/qcat). Once a barcode reaches 100k reads, it will trigger the rest of the ARTIC bioinformatics workflow which will map to the severe acute respiratory syndrome coronavirus isolation from Wuhan, Hubei District, China (Genbank: MN908947.3) using minimap2. This alignment will then be used to generate consensus sequences and variant calls using medaka (https://github.com/nanoporetech/medaka). The entire ONT analysis pipeline is available at https://github.com/gagekmoreno/SARS-CoV-2-in-Southern-Wisconsin.

### Phylogenetic analysis

All 247 available full length sequences from Dane and Milwaukee County through 26 April 2020 were used for phylogenetic analysis using the tools implemented in Nextstrain custom builds (https://github.com/nextstrain/ncov) ^4,52^. Time-resolved and divergence phylogenetic trees were built using the standard Nextstrain tools and scripts ^4,52^. We used custom python scripts to filter and clean metadata.

An additional subsampled global phylogeny using all available sequences in GISAID as of 21 June 2020 were input into the Nextstrain pipeline. A custom ‘Wisconsin’ profile was made to create a Wisconsin-centric subsampled build to include representative sequences. We defined representative sequences as 20 sequences from each US state, and 30 sequences from each country, per month per year. This subsampled global build includes 5,378 sequences or roughly 11% of the total sequences in GISAID as of 21 June 2020. We also ensured that the nearest phylogenetic neighbors of every Dane and Milwaukee County sequence are included, increasing the total to 5,417 sequences. All available Wisconsin sequences available on GISAID by 21 June 2020 were incorporated. An additional 20 sequences from each US state, and 30 sequences from each county, per month per year, were added. All of the Wisconsin sequences included in this study are listed in the `include.txt` to ensure they were represented in the global phylogeny. The scripts and output are available at https://github.com/gagekmoreno/SARS-CoV-2-in-Southern-Wisconsin.

### Estimating the number of introductions

To estimate the number of unique introductions into Dane and Milwaukee County we first identified the closest cophenetic match of each Dane and Milwaukee County sequence in the global SARS-CoV-2 phylogenetic trees generated by Dr. Rob Lanfear at the Australian National University. These trees are generated using MAFFT ^53^, FastTree ^54^ and are available at https://github.com/roblanf/sarscov2phylo/. If the closest neighbor had an ambiguous date, the next closest was chosen. Any sequences which were not already in the down-sampled alignment described above were added using MAFFT. IQ-TREE ^55^ with 1000 Ultrafast bootstrap replicates ^56^ using the flags `-nt 4 -ninit 10 -me 0.05 -bb 1000 -wbtl -czb`. The clock rate of the maximum likelihood tree was estimated using TreeTime ^52^. We first pruned tips which failed the clock filter (`n_iqd = 4`) and then ran TreeTime with the flags The number of introductions into each region was estimated using the maximum likelihood tree as well as 100 of the bootstrap replicate trees. For each, we first generated a time aligned tree with TreeTime with the flags `infer_gtr=True max_iter=2 branch_length_mode=‘auto’ resolve_polytomies=False time_marginal=‘assign’ vary_rate=0.0004 fixed_clock_rate=0.0008` ^57^. Tips which failed the clock filter were pruned from each tree prior to running TreeTime. The 90% highest posterior region was used to calculate a confidence interval for the time of each node. Next, tips in the tree were assigned to either Dane County, Milwaukee County, the U.S. states, or their country of origin and the ancestral states of nodes in the tree were estimated using TreeTime. A sampling bias correction of 2.5 was used to account for under sampling. Nodes were assigned to the region with the highest assigned probability from TreeTime. For each sample from Dane and Milwaukee county we identified the earliest (in calendar time) node assigned to Wisconsin (Dane County, Milwaukee County, and other Wisconsin) in the path between that tip and the root of the tree. Introduction into Dane and MIlwaukee County is assumed to occur at the time between these nodes and their parent node. As we do not know whether Wisconsin samples included in the tree from other studies are from Dane or Milwaukee County (or elsewhere in Wisconsin), our estimates for the timing of introduction into each county represent the timing of introduction of that lineage into Wisconsin as a whole. The time of introduction was evaluated using the mean estimate as well as the lower and upper limits of the timing for each node. Thus, each bootstrap replicate contributes three lines to the plots shown in **Fig 3B** and **Fig 3C**. Furthermore, our estimates of the number of introductions will be conservative in the case of reimportations into Dane or Milwaukee County. Because polytomies were not resolved, any Dane or Milwaukee County tips or lineages directly descending from a polytomy were attributed to a single importation event – to the earliest Wisconsin node.

We also conducted a rarefaction analysis to assess the impact of sampling within Dane and Milwaukee County on the estimated number of introductions. This was done using the time aligned maximum likelihood tree described above. N (20 to 240, in increments of 20) sequences were randomly sampled from the set of Dane and Milwaukee County sequences and all non-sampled Dane and Milwaukee County sequences were pruned from the tree prior to ancestral state reconstruction and estimation of the number of introductions as described above. Ten replicates for each N were conducted.

Code to replicate this analysis is available at https://github.com/gagekmoreno/SARS-CoV-2-in-Southern-Wisconsin. Results were visualized using Matplotlib ^58^, Seaborn (https://github.com/mwaskom/seaborn), and Baltic (https://github.com/evogytis/baltic).

### Phylodynamic analysis

Bayesian phylogenetic inference and dynamic modelling were performed with BEAST2 software (v2.6.2) ^59^ and the PhyDyn package (v1.3.6) ^14^. The phylodynamic analysis infers SARS-CoV-2 phylogenies of sequences within a region of interest and exogenous sequences representing the global phylogeny, and uses tree topology to inform a SEIJR compartmental model. For the Bayesian phylogenetic analysis, an HKY substitution model (gamma count=4; K lognormal prior (μ=1, S=1.25)) and a strict molecular clock (uniform prior 0.0005 to 0.005 substitution/site/year) were used. To select the exogenous sequences, a maximum-likelihood global phylogeny was generated with IQTree and randomly downsampled in a time-stratified manner by collection week. Closest cophenetic neighbors for each of the Wisconsin sequences were additionally included, if not present already. Only sequences with coverage of the entire coding region and less than 1% of N base calls were used. For the Dane County analyses, 107 local and 107 exogenous SARS-CoV-2 sequences were used. For the Milwaukee County analyses, 117 local and 129 exogenous SARS-CoV-2 sequences were used.

The SEIJR model dynamics are defined by the following ordinary differential equations:

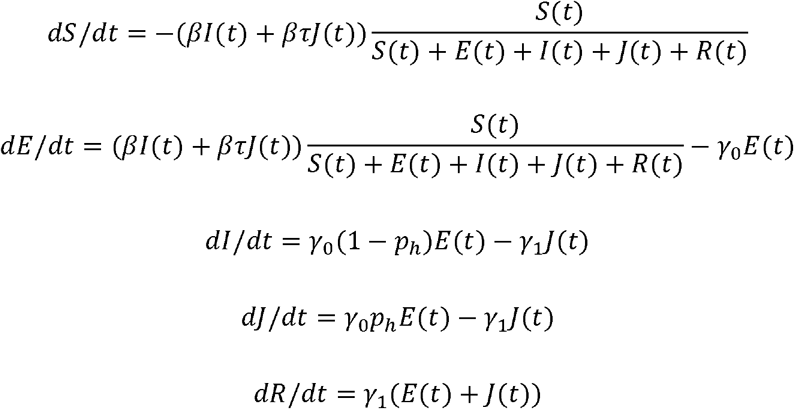

The dynamics of the exogenous compartment is defined by:

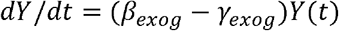

During phylodynamic model fitting, *β, β_exog_*, and *α* are estimated. Estimated R_0_ was derived from *β* as follows.

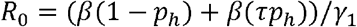

The SEIJR model includes a ‘high transmission’ compartment (J) that accounts for heterogeneous transmission due to superspreading, an important component of SARS-CoV-2 epidemiology ^9,60–62^. Published empirical estimates informed parameterization of superspreading and other epidemiological parameters. The mean duration of latent (1/*γ*_O_) and infectious periods (1/*γ*_1_) was 3 and 5.5 days, respectively ^63^. Likewise, the mean duration of infection for the exogenous compartment (1/*γ*_*exog*_) was fixed at 8.5 days. To model low, medium, and high transmission heterogeneity, the proportion of infectious individuals in the J compartment (*p*_*h*_) and their transmission rate multiplier (τ) were set to 0.2 and 16, 0.1 and 36, or 0.05 and 76, respectively. These *p*_*h*_ and *τ* settings result in 20, 10, or 5% of individuals contributing 80% of total infections. The initial size of the S compartment was fixed at 5 × 10^5^ for Dane County and 9.5 × 10^5^ for Milwaukee County. To account for changes in epidemic dynamics after the Executive Orders, a 25% reduction in importation/exportation of sequences was applied at a 25 March breakpoint, per observed reductions in Google mobility indices for individuals in Wisconsin ^64^. Additionally, the estimated R_0_ after 25 March was allowed to vary from the pre-intervention R_0_ proportionally by a modifier variable, *α*.

Each analysis was run in duplicate for at least 3 million states in BEAST2. Parameter traces were visually inspected for adequate mixing and convergence in Tracer (v1.7.1). Log files from duplicate runs were merged with LogCombiner and 10% burn-in applied. Similarly, trajectory files from duplicate runs were merged with an in-house R script and 10% burn-in applied. BEAST2 XML files and scripts for exogenous sequence selection and phylodynamic data analysis/visualization are provided in the GitHub repository listed below.

## Data Availability

Sequencing data after mapping to SARS-CoV-2 reference genome (Genbank: MN908947.3) have been deposited in the Sequence Read Archive (SRA) under bioproject PRJNA614504. Derived data, analysis pipelines, and figures have been made available for easy replication of these results at a publicly-accessible GitHub repository: https://github.com/gagekmoreno/SARS-CoV-2-in-Southern-Wisconsin.

All sequences have been uploaded to GISAID and GenBank. Associated accession numbers can be found in supplementary information 1 file.

https://github.com/gagekmoreno/SARS-CoV-2-in-Southern-Wisconsin

## Data availability

Sequencing data after mapping to SARS-CoV-2 reference genome (Genbank: MN908947.3) have been deposited in the Sequence Read Archive (SRA) under bioproject PRJNA614504. Derived data, analysis pipelines, and figures have been made available for easy replication of these results at a publically-accessible GitHub repository: https://github.com/gagekmoreno/SARS-CoV-2-in-Southern-Wisconsin.

## Acknowledgements

We gratefully acknowledge Dr. Trevor Bedford and the entire Nextstrain team for making Nextstrain phylogenetic tools publicly available and for their commitment to tracking the global spread of SARS-CoV-2. We also acknowledge the GISAID team for maintaining the largest public repository of SARS-CoV-2 sequence- and metadata. Lastly, we thank Dr. Louise Moncla for her careful reading of and insightful comments on this manuscript.

This project was funded in part through a COVID-19 Response grant from the Wisconsin Partnership Program at the University of Wisconsin School of Medicine and Public Health. KMB is supported by F30 AI145182-01A1 from the National Institutes of Allergy and Infectious Disease. GKM is supported by an NLM training grant to the Computation and Informatics in Biology and Medicine Training Program (NLM 5T15LM007359).

## Supplemental Figures

**Supplemental Figure 1.**
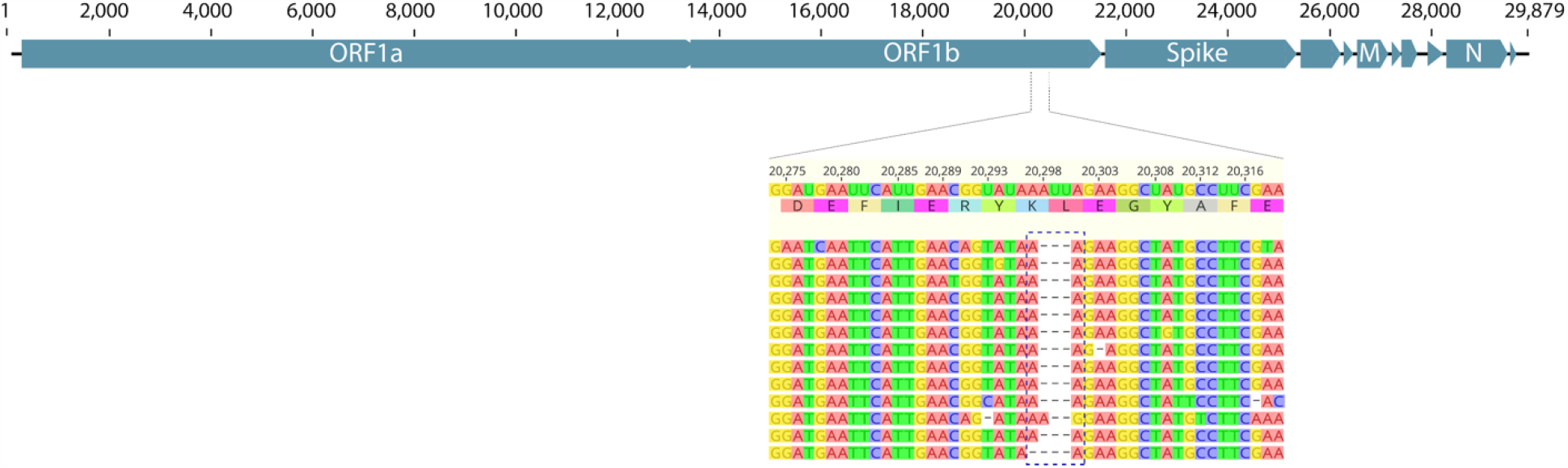
Diagnostic deletion in the index Dane County sample. Consensus-level deletion identified in the Dane County index sample. Zoomed in panel shows nucleotide and amino acid identities of the in-frame deletion.

**Supplemental Figure 2.**
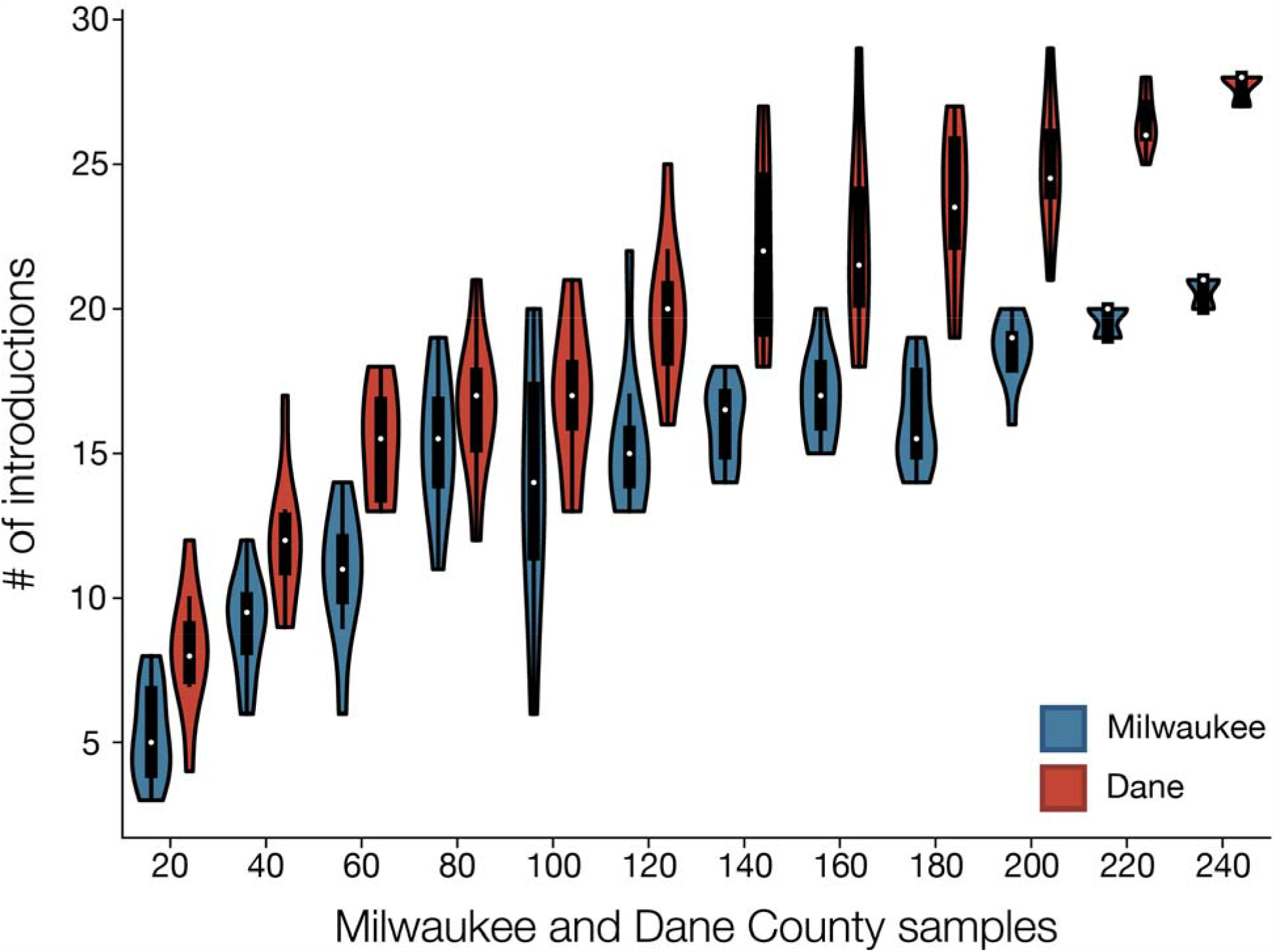
Sampling sensitivity of estimates for the number of introductions into Dane and Milwaukee Counties. Estimates of the number of introductions into Dane and Milwaukee Counties using a time aligned maximum likelihood phylogeny. N sequences (x-axis) were randomly sampled from the available Dane and Milwaukee County samples and the remaining were pruned from the tree. Ten replicates of each N were conducted and the number of introductions (y-axis) was estimated for each.

